# Early Lung Cancer Detection Using Nucleotide Transition Probabilities in plasma cell-free DNA

**DOI:** 10.1101/2025.09.09.25335450

**Authors:** Jinwen Ji, Ruyue Xue, Xu Zhang, Meijia Yang, Lifeng Li, Xiaoran Duan, Wanglong Deng, Rui Yan, Zhihui Xu, Cong Pian, Jie Zhao

## Abstract

Lung cancer, the most lethal malignancy globally, urgently requires effective early detection methods. Current non-invasive approaches based on plasma cell-free DNA (cfDNA) fragmentomics are often constrained by limited sensitivity in early-stage patients due to low tumor DNA fraction. To overcome this, we introduce a novel computational feature—First-Order Transition Probability (FOTP)—to decode nucleotide sequential dependencies within cfDNA fragments. Through systematic analysis of 1,036 participants and low-pass whole-genome sequencing, we demonstrate that the first 10 bp at the 5′ end harbor the most discriminative information for cancer detection. An SVM model leveraging FOTP achieved an AUC of 0.942, with 73.9% sensitivity for stage I and 81.8% for stage II lung cancer at 95% specificity, significantly outperforming existing fragmentomic features. Furthermore, the method generalized robustly across independent and multi-cancer validation sets, including HCC, CRC, and HNSCC, and exhibited potential for tissue-of-origin identification. These findings are supported by nucleotide frequency stability and entropy patterns beyond the initial 10 bp, reflecting underlying nuclease cleavage biases and chromatin features. This work establishes FOTP as a biologically interpretable and highly efficient feature for pan-cancer early detection, offering a scalable pathway toward population-wide screening programs.

## Introduction

Lung cancer is often detected at advanced stages, with the 5-year survival rate being less than 10% for stage III and IV patients [1, 2]. Although low dose computed tomography (LDCT) has been shown to reduce mortality in high-risk (HR) populations, such as long-term smokers, screening uptake remains low due to concerns about radiation exposure and the absence of early symptoms [3]. These challenges underscore the urgent need for a rapid, accurate, and accessible method for early screening.

In recent years, non-invasive cancer detection through liquid biopsy has gained significant attention [4–11]. Among the various plasma components, circulating cell-free DNA (cfDNA) has emerged as a promising biomarker for early cancer diagnosis [12]. cfDNA consists of fragments released from apoptotic and necrotic cells, carrying epigenetic and genomic signatures from their tissues of origin [13, 14]. To date, the detection of early-stage cancer using methylation alterations has mainly been tested with customized panels. Two multi-cancer early detection research have demonstrated that machine learning combined with methylation features can identify tumor-derived signals and determine their tissue of origin [15, 16]. However, panels designed based on prior knowledge and limited genomic regions may fail to capture sufficient tumor signals. For instance, when the fraction of stage I lung cancer–derived DNA in circulation is extremely low (ranging from 0.06% to 0.18%), the median sensitivity drops to 30% [3, 15–17]

Previous studies have shown that genome-wide characteristics may be more sensitive to subtle variations in cfDNA. Analyses of mutational signatures extracted from low-coverage whole-genome sequencing (WGS) data have revealed that informative passenger mutations have the potential to enhance early-stage cancer diagnosis [18]. Although multiple filtering steps are applied to remove germline mutations and sequencing artifacts, concerns regarding the reproducibility of these signatures remain. In contrast, fragmentation profiles generated by enzymatic degradation processes have demonstrated high consistency even in ultra-low-pass data. To improve practical applicability, researchers have partitioned the genome into 5-megabase (Mb) regions and examined alterations in the ratio of short to long fragments for cancer detection [9]. This approach successfully identified approximately 70% of early-stage cancer cases with 98% specificity. Several features closely associated with chromatin accessibility—such as mean coverage, Fast Fourier Transform (FFT)-based features at transcription factor binding sites [18], and the information-weighted fraction of aberrant fragments [10]—have not only exhibited high performance in distinguishing cancer from healthy individuals but also shown potential for inferring tissue of origin. These performance levels remained robust even at 0.1× coverage [18]. Moreover, the diversity of 4 bp end motif at preferential genomic ends was found to be higher in hepatocellular carcinoma (HCC) samples compared with hepatitis B virus (HBV) and healthy controls [19, 20]. When integrated with machine learning models, the end-motif and its analogous 6 bp breakpoint motif achieved high sensitivity in detecting early-stage lung cancer [21–23]. A recent large-scale retrospective study of the Jinling cohort revealed that fragmentomics-based models exhibited robust performance in multi-cancer detection, with an overall sensitivity of 53.5% and a specificity of 98.1% [24].

There have been reports suggesting that adjacent nucleotide dependence can be effectively captured using transition probabilities [25–29]. Unlike direct frequency-based methods, transition probabilities remain robust as sequence length increases, avoiding sparse matrices caused by low sequencing depth, and are suitable for analyzing longer 5′ end sequences of cfDNA fragments. In this study, we hypothesized that transition probabilities at plasma cfDNA fragment ends could serve as biomarkers for early cancer detection. To systematically evaluate the contribution of sequences of different lengths, we derived transition probability features from sequences ranging from 10 bp to 50 bp at the fragment ends. When applied to a predominantly stage I lung cancer dataset, we found that the information embedded within the first ten nucleotides exerted a decisive influence on early-stage cancer detection. Moreover, analysis of an external multi-cancer dataset showed that models based on this method achieved relatively good performance across different cancer types and revealed that different cancer types clustered separately, suggesting that nucleotide transition probability may represent a generalized feature capable of both accurately detecting early-stage cancer and identifying its tissue of origin.

## Method

### Study design

A total of 1,036 participants were enrolled in this prospective observational study from the First Affiliated Hospital of Zhengzhou University (Figure S1, Table 1). The training and testing cohort comprised 332 non-cancer individuals, including 80 with benign lung nodules confirmed by CT or biopsy, and 193 NSCLC patients, of whom approximately 80% were diagnosed at early stages (Stage I–II). Among the NSCLC cases, there were 173 lung adenocarcinoma (LUAD) and 20 lung squamous cell carcinoma (LUSC) cases. Two participants were excluded due to quality control (QC) failure.

**Table 1.**
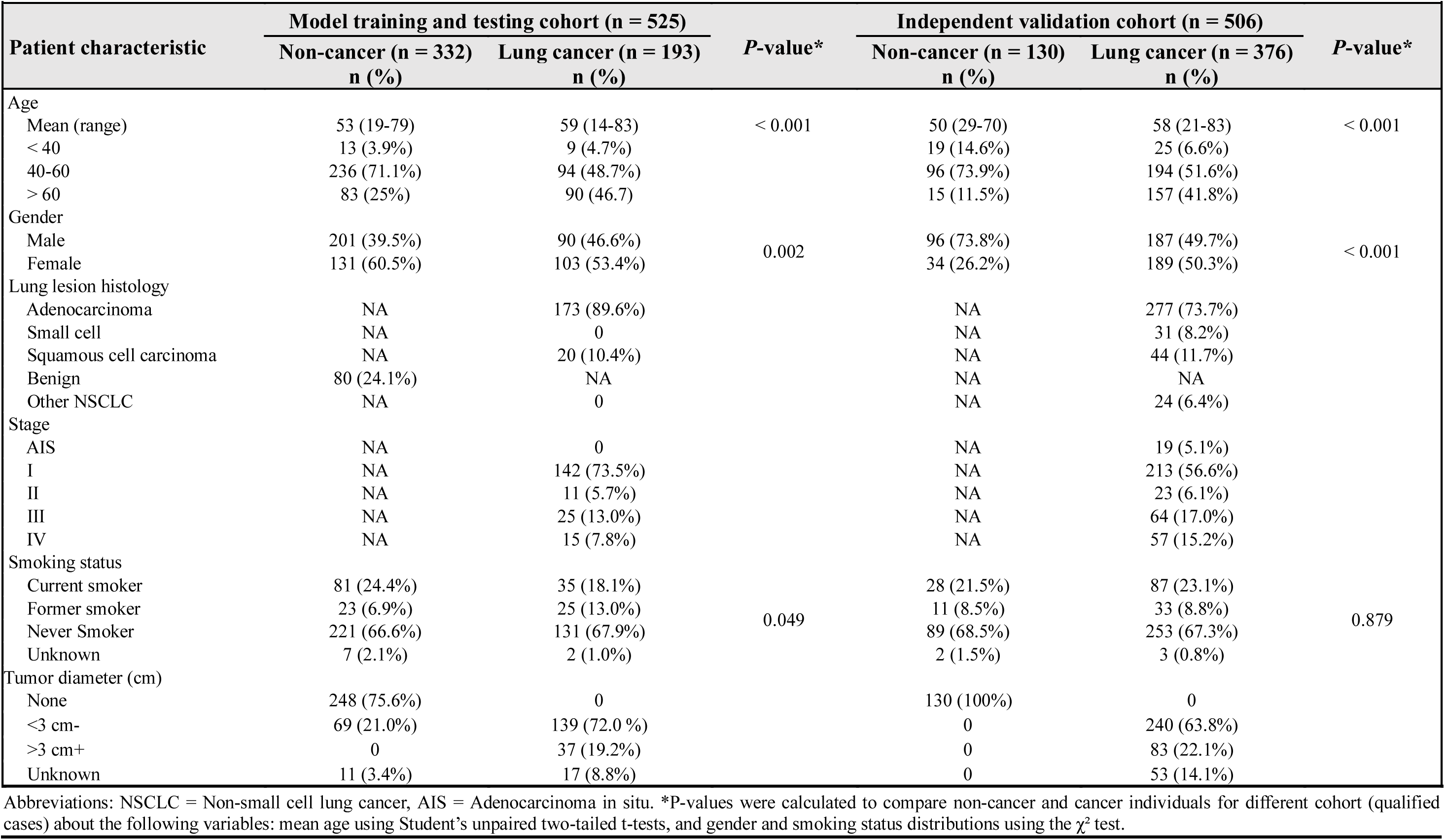
Demographic and clinical characteristics of patients enrolled in this study.

The independent validation cohort consisted of 130 healthy individuals and 376 lung cancer patients, with approximately 70% at early stages, including adenocarcinoma in situ (AIS). Three participants were excluded due to QC failure. Sex was self-reported and verified using X/Y chromosome copy number profiles generated by ichorCNA [30].

To further assess model performance, we obtained 129 publicly available samples [10], comprising 38 healthy individuals, 34 hepatocellular carcinoma (HCC) patients, 17 chronic hepatitis B patients without HCC, and 10 cases each of colorectal cancer (CRC), head and neck squamous cell carcinoma (HNSCC), lung cancer (LC), and nasopharyngeal carcinoma (NPC).

### Sample collection and processing

Plasma samples were collected from healthy individuals, patients with benign pulmonary nodules, and lung cancer patients at the First Affiliated Hospital of Zhengzhou University, prior to tumor resection or therapy. The study was conducted in accordance with the Declaration of Helsinki, with written informed consent obtained from all participants and ethics approval granted by the hospital’s committee (Approval No. 2020-KY-0379-002). Blood samples were centrifuged at 1,600 × g for 10 minutes, followed by 16,000 × g for 10 minutes at 4 °C to remove residual cellular debris. cfDNA was extracted from 10 mL plasma using the QIAamp Circulating Nucleic Acid Kit (Qiagen) and quantified and qualified with a Bioanalyzer 2100 (Agilent Technologies).

### Sequencing Library Preparation

Whole-genome sequencing libraries were prepared from plasma DNA using the TruSeq Nano DNA Library Prep Kit (Illumina). Briefly, 5–10 ng of cfDNA from each sample underwent end-repair, A-tailing, and adapter ligation. Libraries were not subjected to fragment size selection and were sequenced in 150 bp paired-end mode on a NovaSeq 6000 platform to an average depth of ~3×. To minimize bias, case and control samples were matched for sex and age prior to library preparation and sequencing.

### Low-pass whole genome sequencing data processing

FASTQ files were processed using fastp (version 0.22.0) to remove adapters and generate clean reads. The clean reads were aligned to the human reference genome hg19 using BWA (version 0.7.17) with the bwa-mem command and default parameters. Duplicate reads were marked using samblaster (version 0.1.26). Sequencing reads were sorted with samtools (version 1.3.1), and BAM file indices were generated using sambamba (version 0.6.6). BAM files were further filtered with custom Python scripts to remove unmapped reads, low-quality reads, marked duplicates, and read pairs without perfect matches between read1 and read2. Data from previously published studies were processed using the same pipeline.

### Fragmentomic Feature Extraction and Model Development

The hg19 genome was partitioned into non-overlapping 5 Mb bins, from which 473 high-mappability regions were retained [31]. Fragment size ratio (FSR) profiles were generated, and GC-content bias was corrected using LOWESS regression for short (100–150 bp) and long (151–220 bp) fragments [9]. End motif (EDM) and breakpoint motif (BPM) profiles were derived from fragment ends in high-mappability regions [19, 21]. Arm-level copy number variation (CNV) statistics were calculated for 39 non-acrocentric chromosome arms [30].

Principal component analysis (PCA) was applied to the FOTP feature matrix, and the resulting components were used to train support vector machine (SVM) classifiers. Models were trained using 5-fold cross-validation on 80% of the training and testing cohort, with the remaining 20% as a validation set. Performance was evaluated on an internal validation set, an independent validation cohort, and an external cohort. For the small external cohort, SVM models were trained iteratively using a leave-one-out approach.

### Transition Probabilities of cfDNA fragments

For each sample, the transition probabilities of plasma cfDNA fragment end sequences of length N were calculated. The initial probability 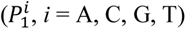 represents the frequency of the four nucleotides across the sequence, and the first-order transition probability 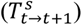 describes nucleotide transitions from position t to t+1 (t = 1,…,N−1), where s denotes one of the 16 possible dinucleotide types (AA, AC, …, TT) (Fig. 1). For each position, the sum of the four transition probabilities per nucleotide group equals 1, as shown in Equation 1.

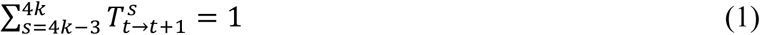

**Fig. 1.**
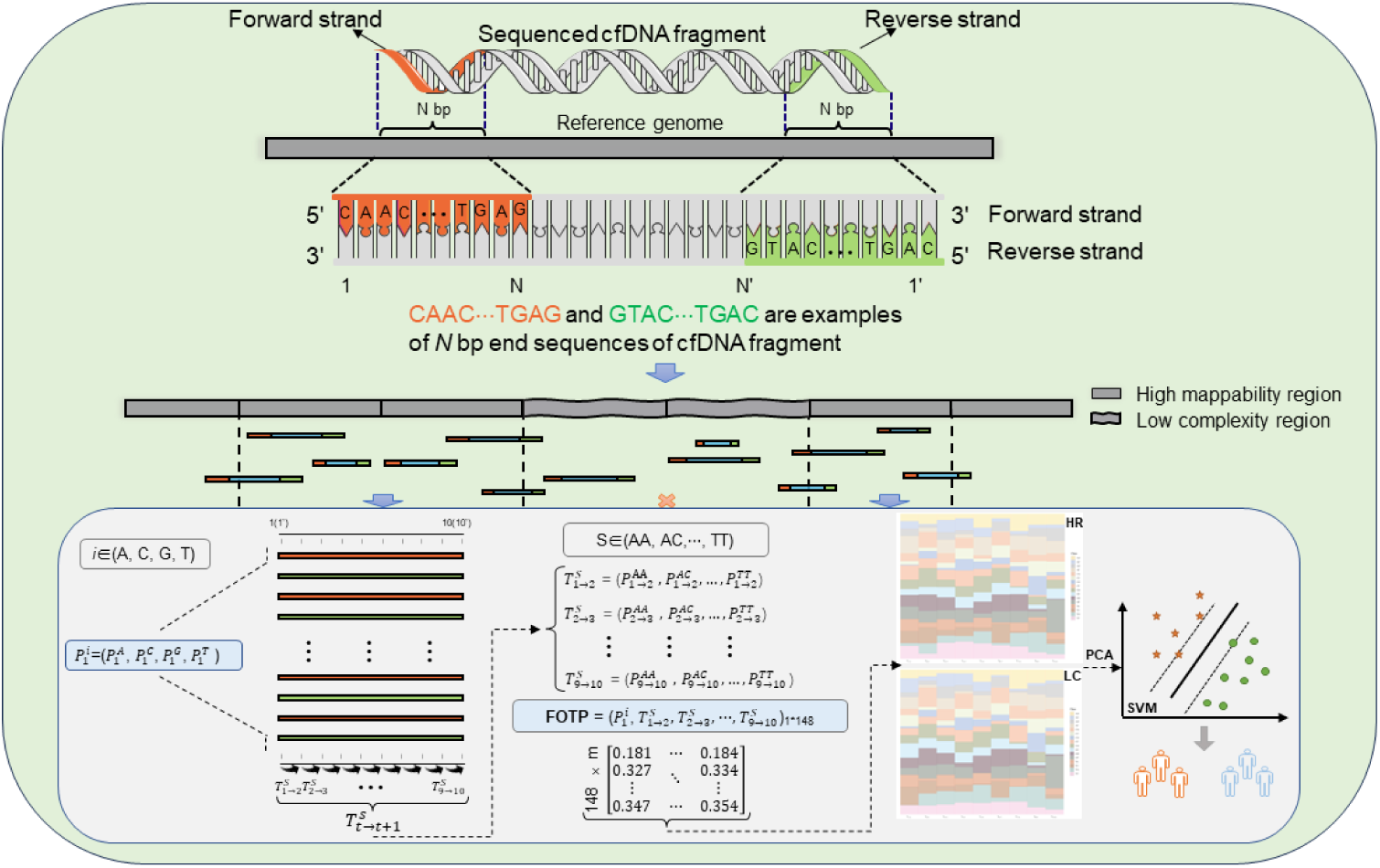
Overview of First-Order Transition Probability (FOTP) Profiling in cfDNA. Only paired-end reads mapped to high-mappability regions of the human reference genome were utilized for FOTP profiling. The initial probability 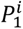 represents the frequency of four nucleotides of the sequence, i ∈ {A, C, G, T}. 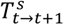 denotes the transition probability from position t to position t+1, with nucleotide dinucleotide state S (e.g., AA, AC, …, TT). FOTP features were profiled for both tumor patients and healthy controls within the cohort. Subsequently, principal component analysis (PCA) was applied for dimensionality reduction, followed by support vector machine (SVM) modeling to evaluate classification performance.

Where k ∈ {1, 2, …, 13}. Combining the initial and transition probabilities yields a vector of 4 + 16 * (N − 1) dimensions.

Second-order transition probabilities (SOTP) are similar to FOTP but are calculated based on 64 trinucleotide types (AAA, AAC, …, TTT), with the probabilities from positions t and t+1 to t+2 summing to 1.

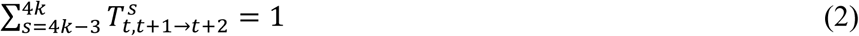

Where k ∈ (1, 2, …, 61). The resulting SOTP vector has 528 dimensions 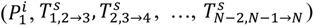, where 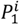 denotes initial dinucleotide probabilities and 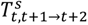 the corresponding transition probabilities.

### Statistical analysis

All statistical analyses were performed in R (version 4.1.1). For subsequent analyses, we employed the output of the SVM decision function, which reflects the signed distance of each sample to the separating hyperplane, as the classification score. Differences in classification scores and representative transition types between groups were evaluated using the Wilcoxon rank-sum test. ROC curves were generated with the pROC package (version 1.18.0). Two-sided P values < 0.05 were considered statistically significant.

## Results

### Position-Specific Nucleotide Transition Probability Profiling

End motif and breakpoint motif have been shown good classification performance for tumor detection [19–21]. To investigate whether longer cfDNA fragment sequences from 5’ ends carry more information, we defined a novel feature representation based on nucleotide transition probabilities, termed FOTP (First-Order Transition Probability). The FOTP feature set incorporates both the initial nucleotide composition and position-specific first-order transition probabilities derived from DNA sequences. For each sample, the initial probabilities correspond to the relative frequencies of the four nucleotides (A, C, G, and T) of the sequence. The transition probabilities are computed for each consecutive pair of positions (t, t+1) along the sequence. For each position t, the probability of transitioning from a given nucleotide at position t to each of the four nucleotides at position t+1 is calculated, yielding 16 possible dinucleotide transitions per position. The probabilities are normalized for each position t, such that the sum of the four outgoing transition probabilities from any given nucleotide equals 1 (Fig. 1). For example, in a sequence of length 10 base pairs, there are 9 distinct transition steps, resulting in 144 transition probability features. Combined with the 4 initial probability features, this produces a 148-dimensional feature vector for each sequence. This approach limits the growth of feature dimensionality, enabling the assessment of sequence composition differences between tumor-derived and healthy cfDNA across extended sequence contexts.

### First 10 bp of cfDNA Fragments Are Critical for Lung Cancer Detection

Given that the majority of cfDNA fragments exceed 100 base pairs in length, we restricted the analysis to the first 50 bp at the 5′ ends to reduce feature redundancy caused by sequence repetition. FOTP features were then calculated from consecutive 10 bp windows within this region and used to train a support vector machine (SVM) model. After evaluation via 5-fold cross-validation and an internal test set, we found the model utilizing features derived from the first 10 bp at the 5′ end achieved the highest performance, yielding an area under the curve (AUC) of 0.938 (95% CI: 0.937–0.938) during cross-validation and 0.942 (95% CI: 0.927–0.956) in the internal test set, with a sensitivity of 77.7% at 95% specificity (Fig. 2A, 2B, Table S1). Furthermore, sensitivity increased with advancing cancer stage, ranging from 74% (95% CI: 67%–81%) in stage I, to 82% (95% CI: 60%–100%) in stage II, 88% (95% CI: 75%–100%) in stage III, and 93% (95% CI: 81%–100%) in stage IV (Table S1).

**Fig. 2.**
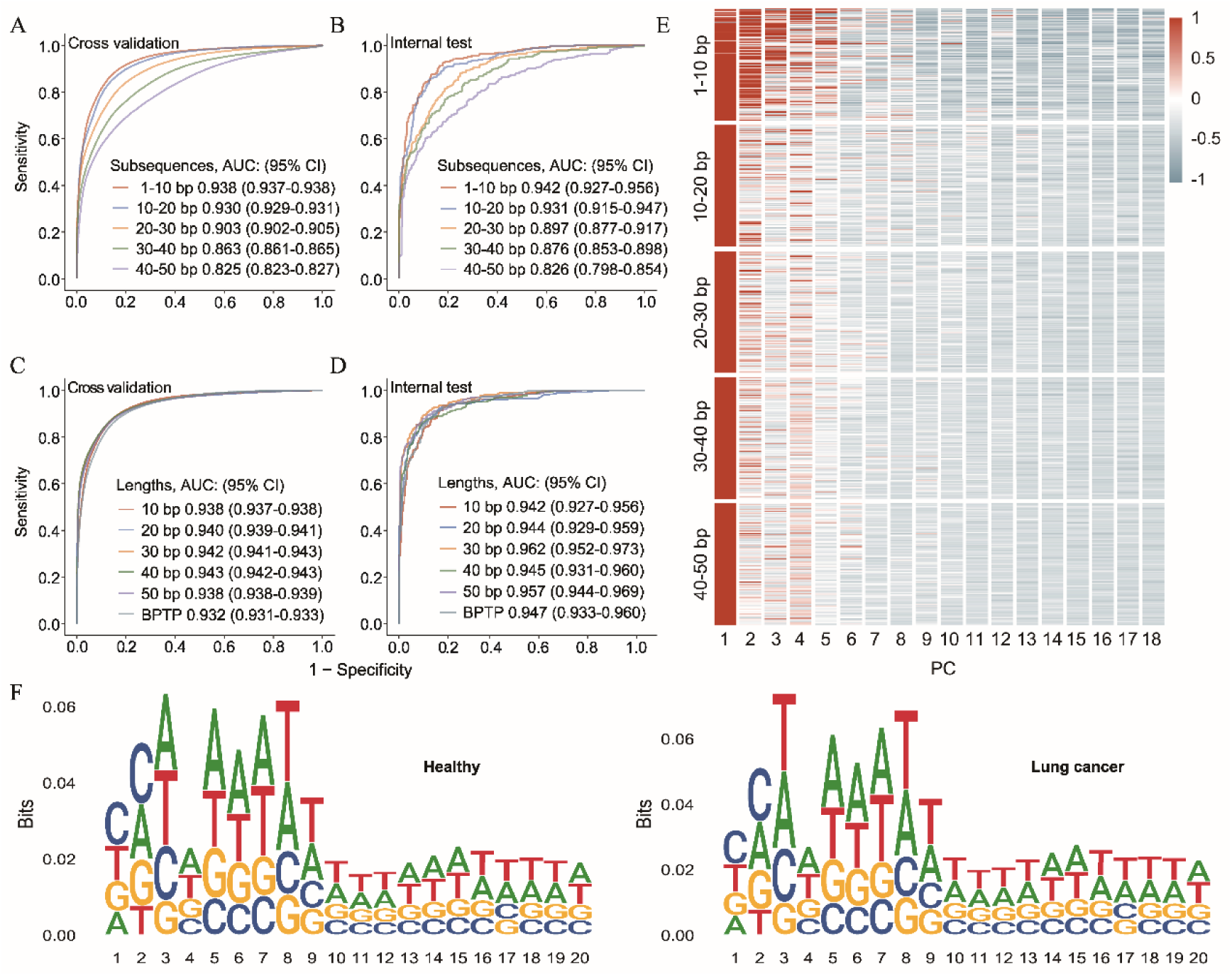
Transition probability from the first ten nucleotides provides the most distinctive information. (A) ROC curves showing the performance of FOTP-based model from sequences at different positions in distinguishing lung cancer patients from healthy individuals in the cross-validation set, and (B) in the internal test set.(C) ROC curves of FOTP derived from sequences of various lengths for differentiating lung cancer patients from healthy individuals in the cross-validation set, and (D) in the internal test set.(E) Principal component loadings of FOTP.(F) Nucleotide frequency within 20 bp from the fragment ends for a healthy individual (left) and a lung cancer patient (right), respectively. Position 1 corresponds to the first base at the 5′ end of each fragment.

Features from 10 bp windows located farther from the 5′ end showed lower performance but still retained some classification ability (Fig. 2A, 2B). Therefore, we assessed whether features from longer sequences (1–10 bp up to 1–50 bp) could provide additional information to improve classification. Due to the superior performance of breakpoint motif–based models [21], we extracted sequences flanking the 5′ fragment ends (10 bp upstream and downstream) to calculate breakpoint transition probability (BPTP) features for comparison with FOTP. Our results showed no significant difference in classification performance between models based on FOTP features across varying sequence lengths and models utilizing BPTP features (Fig. 2C, 2D). Moreover, sensitivity at 95% specificity did not improve with increasing sequence length, and BPTP-based sensitivity decreased by over 10% (Table S2). These results confirm that the first 10 bp at the 5′ end carry the most informative signal for lung cancer detection.

To elucidate these findings, we conducted principal component analysis (PCA) on the features and observed that approximately 20 components captured over 99% of the total variance, irrespective of sequence length (Table S3). The first five principal components explained most of the variance across all sequence lengths (Fig. 2E). These findings corroborated that extending the 5′ end sequence length beyond the initial 10 bp does not provide additional discriminative information.

We further examined the nucleotide composition at positions 1–20 of the 5′ ends of cfDNA fragments in representative cases and controls. In both sample groups, frequencies beyond the first 10 bp were relatively stable, whereas the initial 10 bp exhibited distinct patterns (Fig. 2F and Fig. S1). Position-wise Shannon entropy analysis also confirmed that the variation in Shannon entropy within the first 10 bp was significantly greater than that of the subsequent 10 bp (Fig. S2). In addition, in addition, we observed differences in base composition frequencies between lung cancer and controls, with cancer-derived fragments enriched for cytosine at the first two positions (CC), consistent with DNASE1L3 cleavage specificity[32].

### FOTP-Based Model Outperforms Models Using Other Fragmentomic Features

To evaluate the performance of FOTP, we compared it with several established fragmentomic features of plasma cfDNA in the cohort, including breakpoint motif (BPM), end motif (EDM), copy number variation (CNV), and fragment short-to-long ratio (FSR). The FOTP-based model outperformed other feature-based models in both 5-fold cross-validation and the independent test set (Fig. 3A, B). At 95% specificity, it achieved a median sensitivity of 77.7%, significantly exceeding those of models based on BPM (57.5%), EDM (59.1%), CNV (46.1%), and FSR (50.8%) (Fig. 3C). Sensitivity was higher for the FOTP model than others across all stages except stage II, with a notably high sensitivity of 73.9% in stage I (Fig. 3D), suggesting that FOTP could effectively captures subtle fragmentomic changes of early lung cancer.

**Fig. 3.**
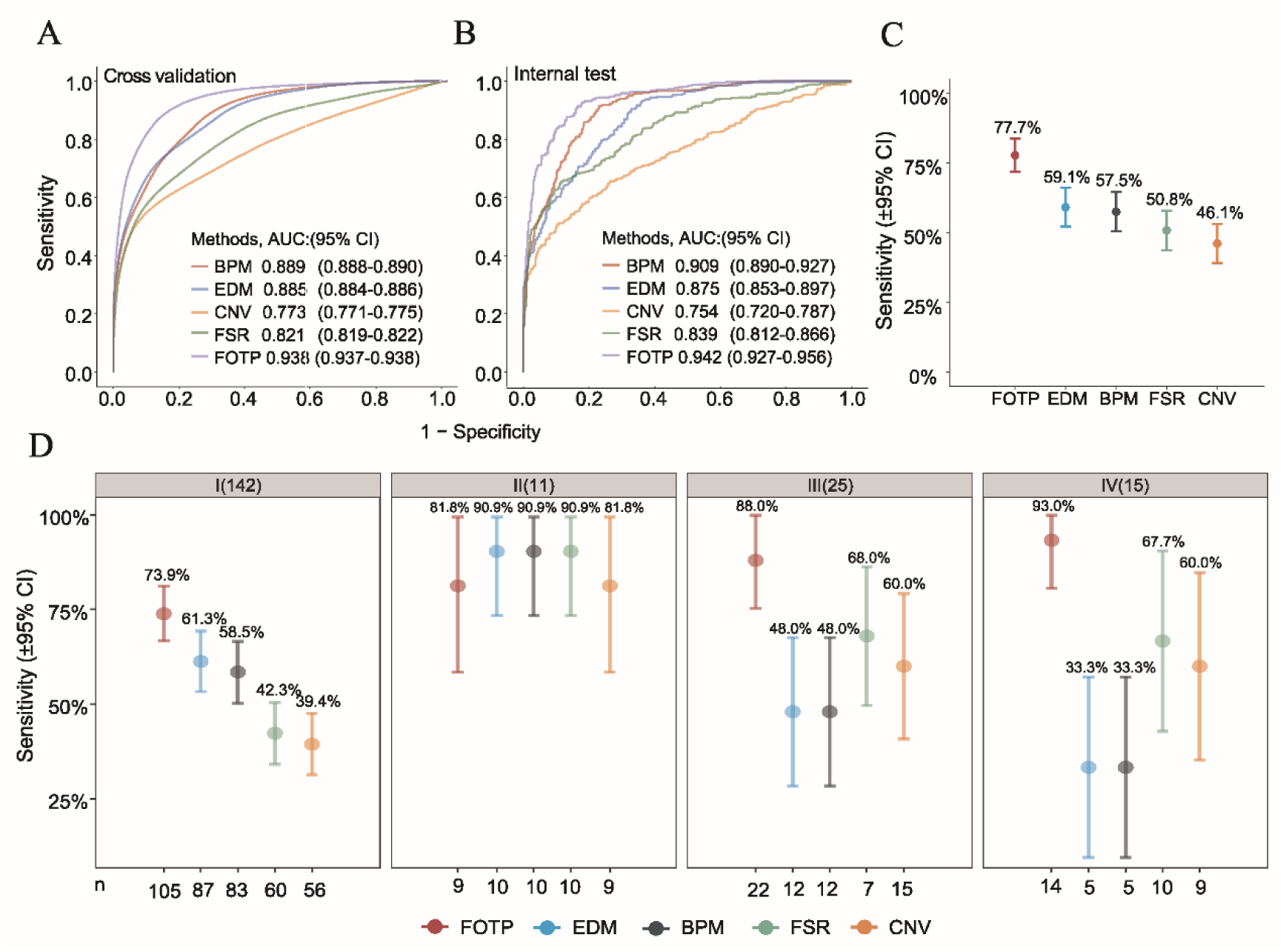
Comparative performance of FOTP and other fragmentomic models. (A) ROC curves comparing models using different fragmentomic features in the cross-validation set, and (B) in the internal test set. (C) Sensitivity of fragmentomic feature-based models at 95% specificity. The X-axis shows the number of correctly predicted cancer samples, and error bars indicate 95% confidence intervals (CI). (D) Sensitivity of five fragmentomic feature-based models across cancer stages at 95% specificity. The X-axis shows the number of correctly identified cancer patients, and the shaded area above indicates stage and sample size.

The discriminative power of the FOTP feature was validated by using the classification scores which quantified differences between healthy individuals and cancer patients. Lung cancer patients exhibited significantly elevated Classification scores compared to non-cancer controls, with scores increasing progressively across cancer stages and reaching the highest levels at stage IV. Although individuals with benign lung lesions showed higher Classification scores relative to healthy controls without cancer, their scores remained significantly lower than those observed in stage I tumor patients, indicating that FOTP effectively differentiates malignant from benign cases (Fig. S4; p < 0.0001, Wilcoxon rank-sum test). These findings demonstrate that Classification scores reflect disease progression and serve as a reliable marker for distinguishing malignancy from benign conditions.

### Evaluation of FOTP-Based Model Performance in an Independent External Cohort

In an independent external cohort of 376 lung cancer patients—68% diagnosed at an early stage—and 130 non-cancer participants, FOTP-base model achieved an AUC of 0.927 (95% CI, 0.904–0.950), slightly lower than in the internal test set (Fig. 4A). Performance remained high across subtypes, with AUCs of 0.911 for LUAD and 0.949 for LUSC. Notably, small-cell lung cancer, which was not included in model training, also exceeded an AUC of 0.95 (Fig. 4B). Sensitivities were 67.1%, 74.0%, 82.8%, and 89.5% for stages I–IV, respectively, and 73.7% for AIS at 95% specificity (Fig. 4C). We analyzed classification scores by cancer stage and subtype, finding significantly higher values in cancer patients (median: AIS = 0.598; stage I = 0.529; stage II = 0.497; stage III = 0.527; stage IV = 0.527) than in healthy individuals (median = 0.168; p < 2.2 × 10^−16^, Wilcoxon rank-sum test) (Fig. S5A). AIS cases were clearly distinguishable from healthy controls, highlighting the high sensitivity of this feature in detecting changes at the earliest stage of lung cancer. In LUAD, scores were more variable with many low values, whereas other subtypes showed consistently high scores, leading to better detection performance (Fig. S5B, Fig. S6A).

**Fig. 4.**
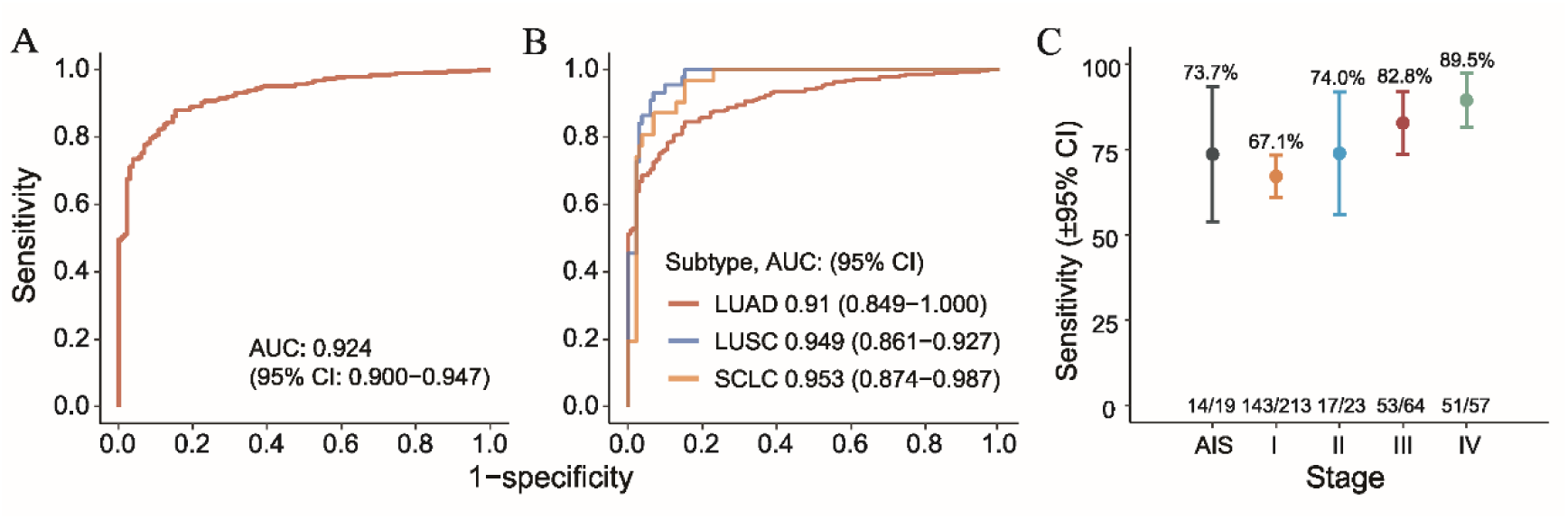
Performance of FOTP-based model in an independent validation cohort. (A) ROC curve showing the overall performance of the FOTP-based model in distinguishing cancer patients from non-cancer individuals in the independent validation cohort. (B) ROC analyses of different lung cancer subtypes in the independent validation cohort. (C) Sensitivity of different cancer stages at 95% specificity. The X-axis indicates the true positive and total cases in each stage. The error bars indicate 95% confidence interval (CI).

We also assessed the influence of demographic and clinical factors on model performance. Age-stratified analysis revealed no significant differences in sensitivity across age groups (Fig. S6B). Model performance was higher in male patients (79.7%) than in female patients (68.3%) (Fig. S6C). Smoking status, known to affect DNA methylation patterns and lung cancer risk, was also reflected in our model’s outputs. Current smokers exhibited higher sensitivity and classification scores, while former and never smokers had lower values (Fig. S6D, S6E). Patients with tumors larger than 3 cm showed higher sensitivity and Classification scores (Fig. S6F, G), likely due to increased tumor cell turnover, resulting in higher release of ctDNA into the blood.

### Application of FOTP to Multi-Cancer Detection

To establish the generalizability of FOTP features beyond lung cancer, we tested the model on a publicly available WGS dataset consisting of 129 plasma DNA samples [19]. The FOTP-based model demonstrated high discriminatory performance among hepatocellular carcinoma (HCC) patients, healthy controls, and hepatitis B virus (HBV) carriers (Fig. 5A). Dimensionality reduction analysis revealed three well-separated clusters corresponding to these groups, with minimal overlap observed between HCC and HBV samples (Fig. 5B). Classification scores were significantly higher in HCC patients compared to healthy individuals (p < 0.0001, Wilcoxon rank-sum test), and effectively differentiated healthy from HBV samples (p < 0.01, Wilcoxon rank-sum test) (Fig. 5C). The multi-cancer classification potential was assessed using five cancer types and non-cancer controls. The model achieved an AUC of 0.931 (95% CI: 0.885–0.977) (Fig. 5D). Classification score distributions clearly distinguished cancer patients from non-cancer individuals (Fig. 5E).

**Fig. 5.**
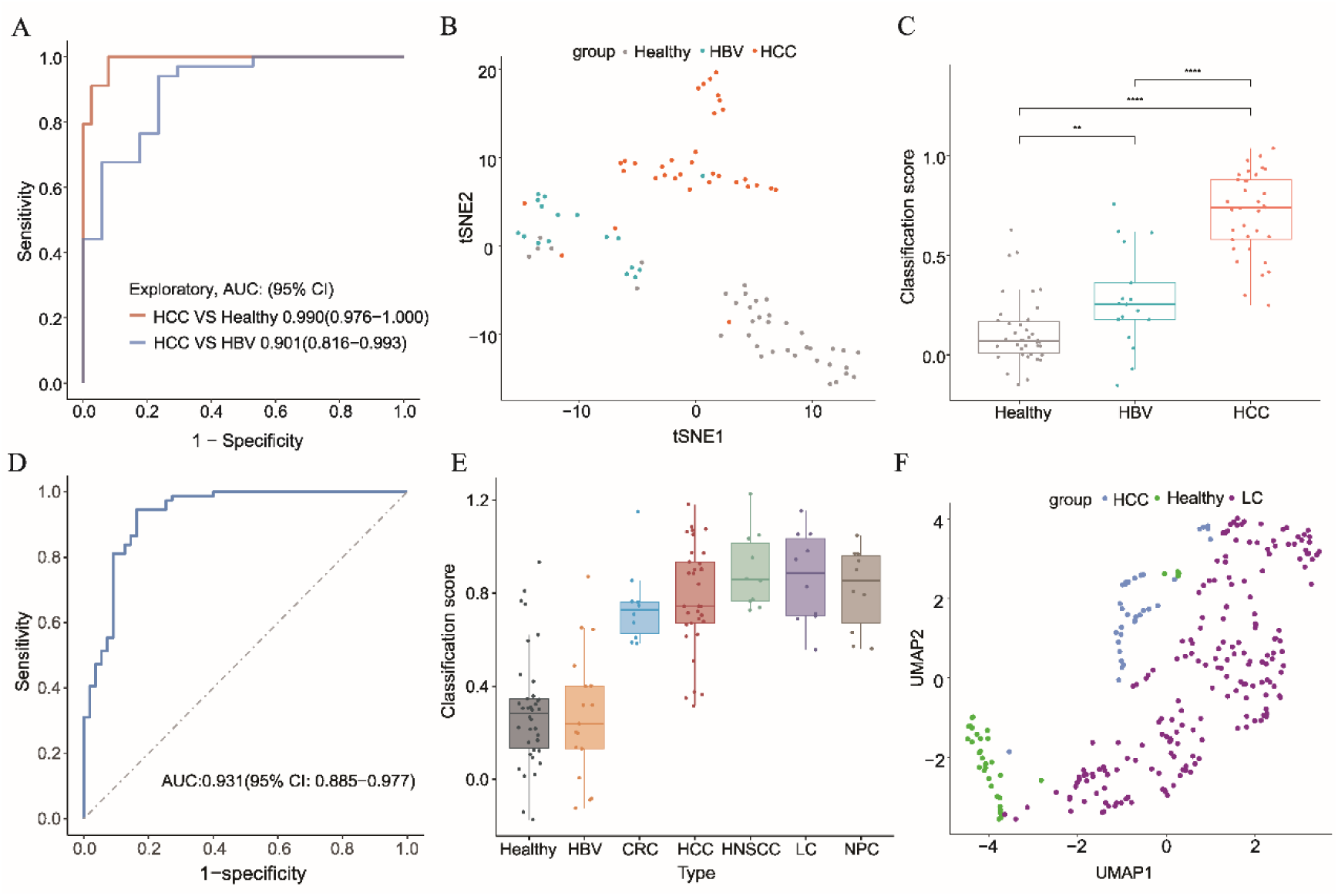
Performance of FOTP-based model in multi-cancer detection. (A) ROC curves for differentiating HCC from non-HCC individuals. (B) t-SNE analysis of HCC patients, HBV patients, and healthy individuals. (C) Classification scores for HCC patients, HBV patients, and healthy individuals. **: p < 0.01, ****: p < 0.0001, Wilcoxon rank-sum test. (D) ROC curves for differentiating mulitple cancer types (HCC, CRC, HNSCC, LC, and NPC). (E) Classification scores of mulitple cancer types in the external dataset. (D) UMAP analysis of HCC patients, and LC patients, and healthy individuals.

To evaluate whether FOTP has potential origin-tracing capability, we performed dimensionality reduction analysis on 34 HCC samples, 38 healthy individuals, and 193 LUAD samples. The results showed that different sample types formed distinct clusters (Fig. 5F), and the substantial differences in nucleotide frequency composition of the first 10 base pairs at the 5′ ends between the two cancer types conferred the capability for origin tracing (Fig. S7).

### SOTP Provides Slightly Improved Performance Over FOTP

Building on the superior performance of FOTP in lung cancer detection, we investigated whether incorporating higher-order nucleotide dependencies could improve classification. Second-order transition probabilities (SOTP) extend FOTP by capturing dependencies among three consecutive nucleotides. Using SOTP features from the first 10 bp of fragment 5′ ends, we extracted 528 features to distinguish lung cancer patients from healthy individuals, achieving AUCs of 0.958 in cross-validation and 0.963 in internal validation set, approximately 2% higher than the FOTP model (Fig. 6A, 6B).

**Fig. 6.**
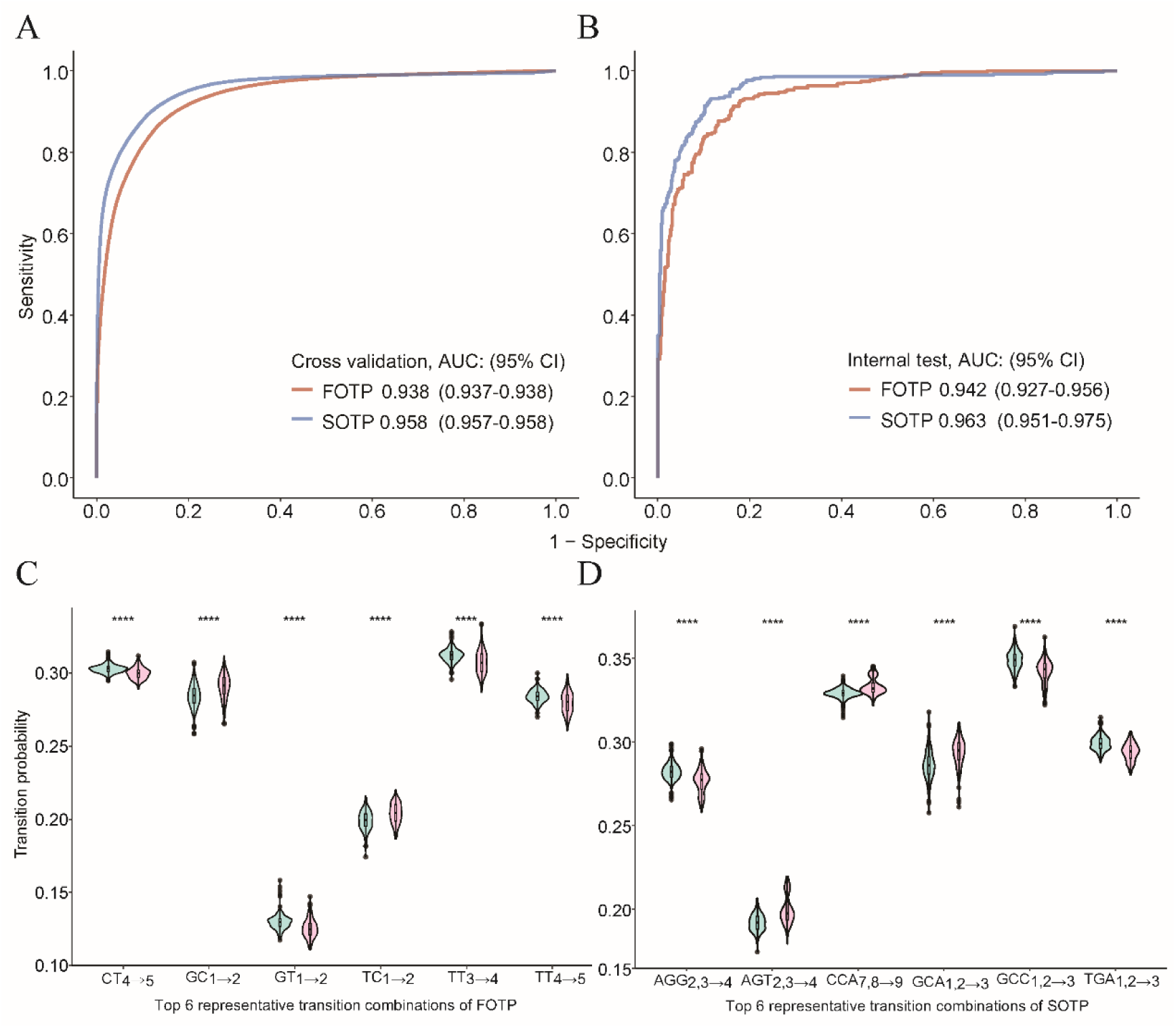
Assessment of the efficacy of FOTP and SOTP in cancer detection. (A) ROC analysis of the performance of FOTP and SOTP for cancer detection in the cross validation and (B) internal test set. (C) The incidence rate of six representative nucleotide combinations of FOTP and (D) SOTP among individuals with or without lung cancer. ****: p < 0.0001, Wilcoxon rank-sum test.

To determine whether specific nucleotide combinations were particularly informative, we analyzed the most discriminatory patterns captured by FOTP and SOTP. In FOTP, two of six representative 2-mer transitions (GC and TC at positions 1–2) were significantly enriched in lung cancer samples, while the others were more enriched in controls (Fig. 6C, p < 0.0001, Wilcoxon rank-sum test). Among the 287 significantly different trinucleotide combinations identified in SOTP, approximately half (n = 154) overlapped with FOTP, yet only two of six representative SOTP combinations (GCA and GCC at positions 1–3) corresponded to the GC1–2 pattern in FOTP and showed opposite trends (Fig. 6D, p < 0.0001, Wilcoxon rank-sum test). These results indicate that SOTP captures distinct, non-redundant sequence context information, enhancing predictive power over FOTP.

## Discussion

cfDNA fragmentation is a non-random process shaped by nucleases with distinct cleavage motifs, such as DNASE1L3 (CCCA) and DNASE1 (TGTG), which have been utilized in end motifs and breakpoint motifs–based methods to distinguish cancer from healthy individuals [12,19–21,23,33–37]. However, fragment cleavage is also shaped by chromatin accessibility, nucleosome positioning, etc. [10,18], which may alter base composition frequency within cfDNA fragment interior, raising the question of whether extensive sequence-level differences exist. Base frequency–based approaches cannot capture such effects, as their dimensionality increases exponentially with sequence length, limiting their ability to evaluate the contribution of longer sequences. Hence, we proposed a novel cfDNA fragmentomic feature that quantifies transition probabilities between adjacent nucleotides at the 5′ ends of cfDNA fragments, and we tested its utility for distinguishing healthy individuals from patients with early-stage lung cancer.

Several studies leveraging transcription factor binding site (TFBS) features have demonstrated comparable performance in cancer detection [37–39]. Motivated by this, we hypothesized that transition probabilities within 10–50 bp sequences might provide additional discriminatory power. Systematic evaluation of positional windows revealed that the first 10 bp yielded the best performance, achieving an AUC of 0.942 in the internal validation set and 77.7% sensitivity at 95% specificity, with strong performance in stage I lung cancer, surpassing several existing fragmentomic methods [9,12,15,19,21]. Nucleotide frequency analysis and performance across 10–50 bp sequences further confirmed that regions beyond the first 10 bp exhibited relatively minor differences between healthy individuals and cancer patients, although they still retained some classification information. Unlike signals from nuclease cleavage, tumor-related heterogeneity—including TF binding, DNA modifications, and other regulatory factors—may dilute local sequence information, which could explain why internal position-based FOTP models generally underperform compared with those derived from the first 10 bp.

Since other sequence features have been applied to multi-cancer detection, we further confirmed the generalizability of FOTP in an independent validation cohort and an external dataset comprising patients with HCC, HBV, LC, HNSCC, and NPC. The FOTP-based model effectively distinguished cancer patients from non-cancer individuals across both cohorts, supporting its potential for broad multi-cancer detection. Moreover, dimensionality reduction revealed that HCC and LC samples formed distinct clusters, indicating a degree of cancer-type specificity. Additionally, inspired by the success of SOTP in predicting DNA methylation sites [28,29], we also evaluated SOTP features, which showed slightly better performance than FOTP, likely due to their ability to capture longer-range dependencies or cleavage-related sequence contexts.

Despite these promising findings, several limitations remain. The study cohort was relatively small and derived from a single clinical center, underscoring the need for validation in larger, multi-institutional populations. Although transition probabilities demonstrated cross-cohort generalizability, their effectiveness in detecting cancer types beyond lung and liver cancer requires further evaluation. In summary, FOTP provides a biologically informed and computationally efficient feature for early cancer detection, with proven utility across cancer stages, cancer subtypes, and cohorts. We will further focus on improving its predictive performance and validating its clinical utility in larger cohorts.

## Conclusions

In brief, our research identified a novel feature called transition probability derived from plasma cfDNA fragments for non-invasive cancer detection. This proof-of-concept paper demonstrated that the internal sequences of cfDNA fragments longer than 10 bp at the 5’ end still carry significant information for early cancer diagnosis. The transition probability feature could yield more valuable information for early cancer detection, as the model exhibits superior performance. The generalization ability of our feature was further confirmed in an independent validation cohort and a public dataset. These demonstrate the promising application of our features for multi-cancer early detection.

## Abbreviations

cfDNA: cell-free DNA
tDNA: circulating tumor DNA
FOTP: first-order transition probability
SOTP: second-order transition probability
AUC: area under the ROC curve
MCED: multi-cancer early detection
LDCT: Low-dose computed tomography
NSCLC: non-small cell lung cancer
WGS: whole-genome sequencing
HCC: Hepatocellular carcinoma
CT: computerized tomography
CRC: colorectal cancer
HNSCC: head and neck squamous cell carcinoma
LC: lung cancer
NPC: nasopharyngeal carcinoma
LOO-CV: leave-one-out cross validation
SVM: support vector machine
PCA: principal component analysis
BPTP: breakpoint transition probability
CI: confidence interval
HBV: chronic hepatitis B
DNASE1L3: deoxyribonuclease 1 like 3
BPM: 6-bp breakpoint-motif
EDM: 4-mer end motif
CNV: copy number variation
FSR: fragment size ratio
t-SNE: t-distributed Stochastic Neighbour Embedding
UMAP: The Uniform Manifold Approximation and Projection

## Acknowledgments

The authors gratefully thank the support provided by Biobank of The First Affiliated Hospital of Zhengzhou University (China Human Genetic Resources Preservation Approval No. [2022]BC0079), and WU JIEPING medical foundation and the State Key Laboratory of Translational Medicine and Innovative Drug Development of Nanjing, Jiangsu Simcere Diagnostics Co., Ltd. We thank individuals from our laboratories and collaborating partners for critical review of this work and China Human Genetic Resources Preservation Approval No. [2023]CJ0513 from the Collaborative Innovation Major Project. This study makes use of sequencing data generated by The Chinese University of Hong Kong (CUHK) Circulating Nucleic Acids Research Group.

## Data accessibility

The data that support the findings of this study are available from the corresponding author *Cong Pian* upon reasonable request.

## Conflicts of interest

The authors have declared that no competing interests exist.

## Funding

This work was supported by the Collaborative Innovation Major Project of Zhengzhou (Grant No. 20XTZX08017); Funding for Scientific Research and Innovation Team of The First Affiliated Hospital of Zhengzhou University (ZYCXTD2023005); The key science and technology program in Henan Province (Grant No. 201400210400); Key scientific research project plan of colleges and universities in Henan Province (Grant No.22A416012); The National Natural Science Foundation of China (Grant No. 82002433, 82203028); The Fundamental Research Funds for the Central Universities [JCQY202108, ZJ22195010 and KYCYXT2022010]; and the Startup Foundation for Advanced Talents at Nanjing Agricultural University [050/804009].

## Authors contributions

JWJ, RYX, and XZ contributed equally to this work. JWJ, ZHX and CP were involved into conceptualization and study design. JWJ, RYX, XZ, MJY, LFL, XRD and RY assembled and integrated sequencing and clinical data. JWJ, RYX, XZ, MJY, LFL, XRD, WLD, and RY contributed to resources and investigation. JWJ, ZHX, and CP performed bioinformatics analyses, statistical analysis and writing-original draft preparation. JWJ conducted validation and visualization. RYX, WLD, ZHX, PC and JZ contributed to project administration. JWJ, ZHX, RYX, XZ, CP and JZ contributed to the revision. JZ and CP contributed to funding acquisition. All authors have read and agreed to the published version of the manuscript.

## Notes

### Competing Interest Statement

The authors have declared no competing interest.

### Author Declarations

The ethics committee of the First Affiliated Hospital of Zhengzhou University gave ethical approval for this work(Approval No. 2020-KY-0379-002).

## References

1. Siegel RL, Miller KD, Fuchs HE, Jemal A. Cancer statistics, 2022. CA Cancer J Clin. 2022; 72:7–33.

2. Oudkerk M, Liu S, Heuvelmans MA, Walter JE, Field JK. Lung cancer LDCT screening and mortality reduction - evidence, pitfalls and future perspectives. Nat Rev Clin Oncol. 2021; 18:135–151.

3. Song P, Wu LR, Yan YH, Zhang JX, Chu T, Kwong LN, et al. Limitations and opportunities of technologies for the analysis of cell-free DNA in cancer diagnostics. Nat Biomed Eng. 2022; 6:232–245.

4. Wan JCM, Massie C, Garcia-Corbacho J, Mouliere F, Brenton JD, Caldas C, et al. Liquid biopsies come of age: towards implementation of circulating tumour DNA. Nat Rev Cancer. 2017; 17:223–238.

5. Diaz LA, Bardelli A. Liquid biopsies: genotyping circulating tumor DNA. J Clin Oncol. 2014; 32:579–586.

6. Thierry AR, Mouliere F, El Messaoudi S, Mollevi C, Lopez-Crapez E, Rolet F, et al: Clinical validation of the detection of KRAS and BRAF mutations from circulating tumor DNA. Nat Med. 2014; 20:430–435.

7. Hao TB, Shi W, Shen XJ, Qi J, Wu XH, Wu Y, et al. Circulating cell-free DNA in serum as a biomarker for diagnosis and prognostic prediction of colorectal cancer. Br J Cancer. 2014; 111:1482–1489.

8. Lebofsky R, Decraene C, Bernard V, Kamal M, Blin A, Leroy Q, et al. Circulating tumor DNA as a non-invasive substitute to metastasis biopsy for tumor genotyping and personalized medicine in a prospective trial across all tumor types. Mol Oncol. 2015; 9:783–790.

9. Cristiano S, Leal A, Phallen J, Fiksel J, Adleff V, Bruhm DC, et al. Genome-wide cell-free DNA fragmentation in patients with cancer. Nature. 2019; 570:385–389.

10. Budhraja KK, McDonald BR, Stephens MD, Contente-Cuomo T, Markus H, Farooq M, et al. Genome-wide analysis of aberrant position and sequence of plasma DNA fragment ends in patients with cancer. Sci Transl Med. 2023;15: eabm6863.

11. Shen SY, Singhania R, Fehringer G, Chakravarthy A, Roehrl MHA, Chadwick D, et al. Sensitive tumour detection and classification using plasma cell-free DNA methylomes. Nature. 2018; 563:579–583.

12. Lo YMD, Han DSC, Jiang P, Chiu RWK. Epigenetics, fragmentomics, and topology of cell-free DNA in liquid biopsies. Science. 2021;372: eaaw3616.

13. Luo H, Wei W, Ye Z, Zheng J, Xu RH. Liquid biopsy of methylation biomarkers in cell-free DNA. Trends Mol Med. 2021; 27:482–500.

14. Gao Q, Zeng Q, Wang Z, Li C, Xu Y, Cui P, et al. Circulating cell-free DNA for cancer early detection. Innovation (Camb). 2022; 3:100259.

15. Klein EA, Richards D, Cohn A, Tummala M, Lapham R, Cosgrove D, et al. Clinical validation of a targeted methylation-based multi-cancer early detection test using an independent validation set. Ann Oncol. 2021; 32:1167–1177.

16. Gao Q, Lin YP, Li BS, Wang GQ, Dong LQ, Shen BY, et al. Unintrusive multi-cancer detection by circulating cell-free DNA methylation sequencing (THUNDER): development and independent validation studies. Ann Oncol. 2023; 34:486–495.

17. Bentley DR, Balasubramanian S, Swerdlow HP, Smith GP, Milton J, Brown CG, et al. Accurate whole human genome sequencing using reversible terminator chemistry. Nature. 2008; 456:53–59.

18. Doebley AL, Ko M, Liao H, Cruikshank AE, Santos K, Kikawa C, et al. A framework for clinical cancer subtyping from nucleosome profiling of cell-free DNA. Nat Commun. 2022; 13:7475.

19. Jiang P, Sun K, Peng W, Cheng SH, Ni M, Yeung PC, et al. Plasma DNA end-motif profiling as a fragmentomic marker in cancer, pregnancy, and transplantation. Cancer Discov. 2020; 10:664–673.

20. Jin C, Liu X, Zheng W, Su L, Liu Y, Guo X, et al. Characterization of fragment sizes, copy number aberrations and 4-mer end motifs in cell-free DNA of hepatocellular carcinoma for enhanced liquid biopsy-based cancer detection. Mol Oncol. 2021; 15:2377–2389.

21. Guo W, Chen X, Liu R, Liang N, Ma Q, Bao H, et al. Sensitive detection of stage I lung adenocarcinoma using plasma cell-free DNA breakpoint motif profiling. EBioMedicine. 2022; 81:104131.

22. Bao H, Wang Z, Ma XJ, Guo W, Zhang XY, Tang WXF, et al. An ultra-sensitive assay using cell-free DNA fragmentomics for multi-cancer early detection. Mol Cancer. 2022; 21:1–7.

23. Wang S, Meng F, Li M, Bao H, Chen X, Zhu M, et al. Multidimensional cell-free DNA fragmentomic assay for detection of early-stage lung cancer. Am J Respir Crit Care Med. 2023; 207:1203–1213.

24. Hua Bao, Shanshan Yang, Xiaoxi Chen, Guangqiang Dong, Yuan Mao, Shuyu Wu, et al. Early detection of multiple cancer types using multidimensional cell-free DNA fragmentomics. Nat Med. 2025; 31:2737–2745

25. Almagor H. A Markov analysis of DNA sequences. J Theor Biol. 1983; 104:633–645.

26. Wren JD, Hildebrand WH, Chandrasekaran S, Melcher U. Markov model recognition and classification of DNA/protein sequences within large text databases. Bioinformatics. 2005; 21:4046–4053.

27. Borodovsky M, McIninch JD, Koonin EV, Rudd KE, Medigue C, Danchin A. Detection of new genes in a bacterial genome using Markov models for three gene classes. Nucleic Acids Res. 1995; 23:3554–3562.

28. Pian C, Zhang G, Li F, Fan X. MM-6mAPred: identifying DNA N6-methyladenine sites based on Markov model. Bioinformatics. 2020; 36:388–392.

29. Yang JL, Lang K, Zhang GL, Fan XD, Chen YY, Pian C, Elofsson A. SOMM4mC: a second-order Markov model for DNA N4-methylcytosine site prediction in six species. Bioinformatics. 2020; 36:4103–4105.

30. Adalsteinsson VA, Ha G, Freeman SS, Choudhury AD, Stover DG, Parsons HA, et al. Scalable whole-exome sequencing of cell-free DNA reveals high concordance with metastatic tumors. Nat Commun. 2017; 8:1324.

31. Mathios D, Johansen JS, Cristiano S, Medina JE, Phallen J, Larsen KR, et al. Detection and characterization of lung cancer using cell-free DNA fragmentomes. Nat Commun. 2021; 12:5060.

32. Serpas L, Chan RWY., Jiang PY., Ni M., Sun K, Rashidfarrokhi A, et al. Dnase1l3 deletion causes aberrations in length and end-motif frequencies in plasma DNA. Proc Natl Acad Sci U S A. 2019; 116:641–649.

33. Chan KCA, Jiang PY, Sun K, Cheng YKY, Tong YK, Cheng SH, et al. Second generation noninvasive fetal genome analysis reveals de novo mutations, single-base parental inheritance, and preferred DNA ends. Proc Natl Acad Sci U S A. 2016;113: E8159–E8168.

34. Jiang P, Sun K, Tong YK, Cheng SH, Cheng THT, Heung MMS, et al. Preferred end coordinates and somatic variants as signatures of circulating tumor DNA associated with hepatocellular carcinoma. Proc Natl Acad Sci U S A. 2018;115: E10925–E10933.

35. Zhou Z, Ma ML, Chan RWY, Lam WKJ, Peng W, Gai W, et al. Fragmentation landscape of cell-free DNA revealed by deconvolutional analysis of end motifs. Proc Natl Acad Sci U S A. 2023;120: e2220982120.

36. Han DSC, Ni M, Chan RWY, Chan VWH, Lui KO, Chiu RWK, Lo YMD. The Biology of Cell-free DNA Fragmentation and the Roles of DNASE1, DNASE1L3, and DFFB. Am J Hum Genet. 2020; 106:202–214.

37. Sun K, Jiang PY, Cheng SH, Cheng THT, Wong J, Wong VWS, et al. Orientation-aware plasma cell-free DNA fragmentation analysis in open chromatin regions informs tissue of origin. Genome Res. 2019; 29:418–427.

38. Lo YMD, Chan KCA, Sun H, Chen EZ, Jiang PY, Lun FMF, et al. Maternal Plasma DNA Sequencing Reveals the Genome-Wide Genetic and Mutational Profile of the Fetus. Sci Transl Med. 2010; 2:61ra91.

39. Foda ZH, Annapragada AV, Boyapati K, Bruhm DC, Vulpescu NA, Medina JE, et al. Detecting Liver Cancer Using Cell-Free DNA Fragmentomes. Cancer Discov. 2023; 13:616–631.

