## Supplemental File for "Early Lung Cancer Detection Using Nucleotide Transition Probabilities in plasma cell-free DNA"

**Supplementary Materials**

**This file includes:**

**Tables S1-S3**

**Fig. S1-S7**

**Supplementary Tables**

**Table S1 Sensitivity of FOTP-based models in consecutive 10-bp windows at 95% specificity**

**Table S2 Model sensitivity based on FOTP features derived from sequences of varying lengths at 95% specificity**

**Table S3** **Number of Principal components explaining 99% of total variance**

**Supplementary Figures**

**Fig. S1 Study design**

**Fig. S2 Nucleotide frequencies within the first 20 bp from fragment ends for a healthy individual and an LC patient**

**Fig. S3 Shannon entropy of the first 20 bp at fragment ends**

**Fig. S4 Classification score distributions across non-cancer individuals and cancer patients**

**Fig. S5 Classification score distributions across non-cancer individuals and cancer patients in the independent external test cohort**

**Fig. S6 Sensitivity of FOTP-based model across different clinical subgroups in the independent cohort at 95% specificity**

**Fig. S7 Nucleotide frequencies within the first 20 bp from fragment ends for healthy individuals and HCC patients.**

**Supplementary Tables**

| **Table S1 Sensitivity of FOTP-based models in consecutive 10-bp windows at 95% specificity** | | | | | |
| --- | --- | --- | --- | --- | --- |
| subsequences | 95% Specificity | Ⅰ | Ⅱ | Ⅲ | Ⅳ |
|  | Sensitivity |  |  |  |  |
| 1-10 bp | 0.777  (0.718-0.836) | 105/142  (0.667-0.812) | 9/11  (0.590-1) | 22/25  (0.753-1) | 14/15  (0.807-1) |
| 10-20 bp | 0.663  (0.596-0.730) | 94/142  (0.584-0.740) | 6/11  (0.251-0.840) | 17/25  (0.497-0.863) | 10/15  (0.428-0.905) |
| 20-30 bp | 0.637  (0.569-0.705) | 91/142  (0.562-0.720) | 9/11  (0.590-1) | 15/25  (0.408-0.792) | 8/15  (0.281-0.786) |
| 30-40 bp | 0.523  (0.453-0.593) | 72/142  (0.425-0.589) | 11/11  (1) | 12/25  (0.284-0.676) | 6/15  (0.152-0.648) |
| 40-50 bp | 0.523  (0.453-0.593) | 76/142  (0.453-0.617) | 8/11  (0.464-0.990) | 10/25  (0.208-0.592) | 7/15  (0.214-0.719) |

| **Table S2 Model sensitivity based on FOTP features derived from sequences of varying lengths at 95% specificity** | | | | | |
| --- | --- | --- | --- | --- | --- |
| lengths | 95% Specificity | Ⅰ | Ⅱ | Ⅲ | Ⅳ |
|  | Sensitivity |  |  |  |  |
| 10 bp | 0.777  (0.718-0.836) | 105/142  (0.667-0.812) | 9/11  (0.590-1) | 22/25  (0.753-1) | 14/15  (0.807-1) |
| 20 bp | 0.767  (0.707-0.827) | 105/142  (0.667-0.812) | 9/11  (0.590, 1) | 20/25  (0.643-0.957) | 13/15  (0.695-1) |
| 30 bp | 0.756  (0.695-0.817) | 103/142  (0.652-0.799) | 8/11  (0.464-0.990) | 22/25  (0.753-1) | 13/15  (0.695-1) |
| 40 bp | 0.731  (0.668-0.794) | 101/142  (0.637-0.786) | 8/11  (0.464-0.990) | 20/25  (0.643-0.957) | 12/15  (0.598-1) |
| 50 bp | 0.782  (0.724-0.840) | 107/142  (0.683-0.824) | 8/11  (0.464-0.990) | 23/25  (0.814-1) | 13/15  (0.695-1) |
| 20 bp BPTP | 0.668  (0.602-0.734) | 90/142  (0.555-0.713) | 7/11  (0.352-0.921) | 20/25  (0.643-0.957) | 12/15  (0.598-1) |

| Table S3 Number of Principal components explaining 99% of total variance | | | | | |
| --- | --- | --- | --- | --- | --- |
| lengths | 10 bp | 20 bp | 30 bp | 40 bp | 50 bp |
| Feature dimension | 148 | 308 | 468 | 628 | 788 |
| Numbers of PC  (99% variance) | 20 | 21 | 20 | 19 | 18 |

**Supplementary Figures**

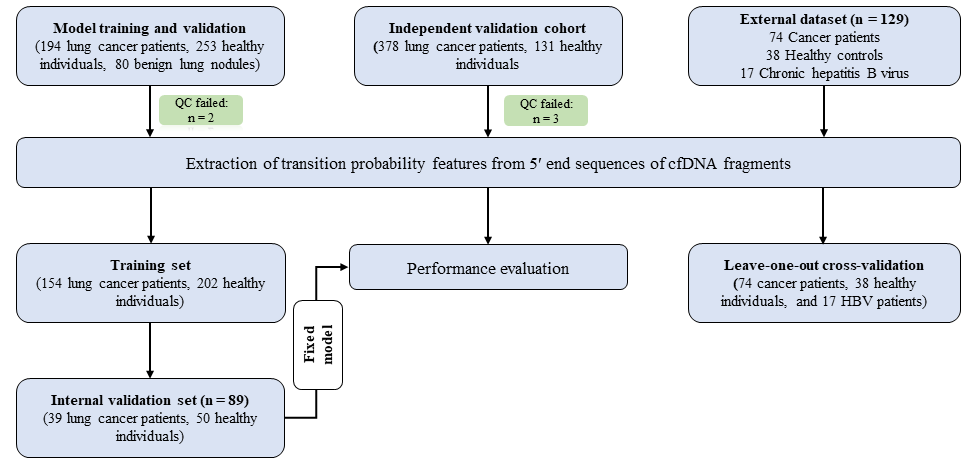

**Fig. S1 Study design**

A total of 1,036 participants were enrolled from the First Affiliated Hospital of Zhengzhou University. Among them, 332 were non-cancer individuals (including 80 with benign lung nodules confirmed by CT or biopsy) and 193 were NSCLC patients. Of these, 525 participants (193 lung cancer patients, 252 high-risk individuals, and 80 with benign lung nodules) were used for model training and performance evaluation. The trained model was subsequently applied to an independent validation cohort comprising 376 lung cancer patients and 130 high-risk individuals to assess its predictive performance. In addition, an external dataset of 129 individuals (38 healthy controls, 17 HBV patients, and 74 cancer patients, including 34 HCC, 10 CRC, 10 HNSCC, 10 LC, and 10 NPC cases) from published studies was analyzed to evaluate the generalizability of our FOTP-based model across other cancer types.

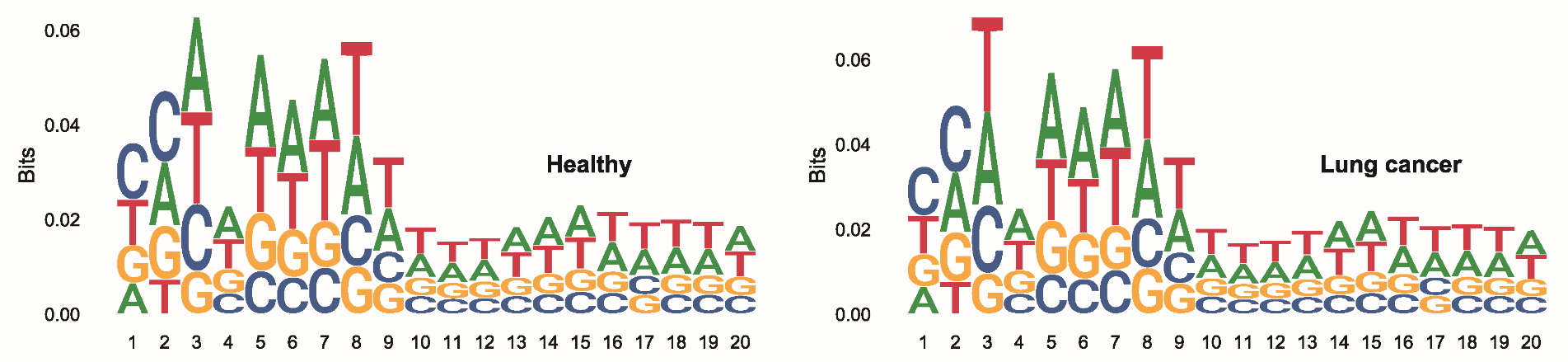

**Fig. S2 Nucleotide frequencies within the first 20 bp from fragment ends for a healthy individual and an LC patient. Position 1 denotes the first base at the 5′ end of fragments.**

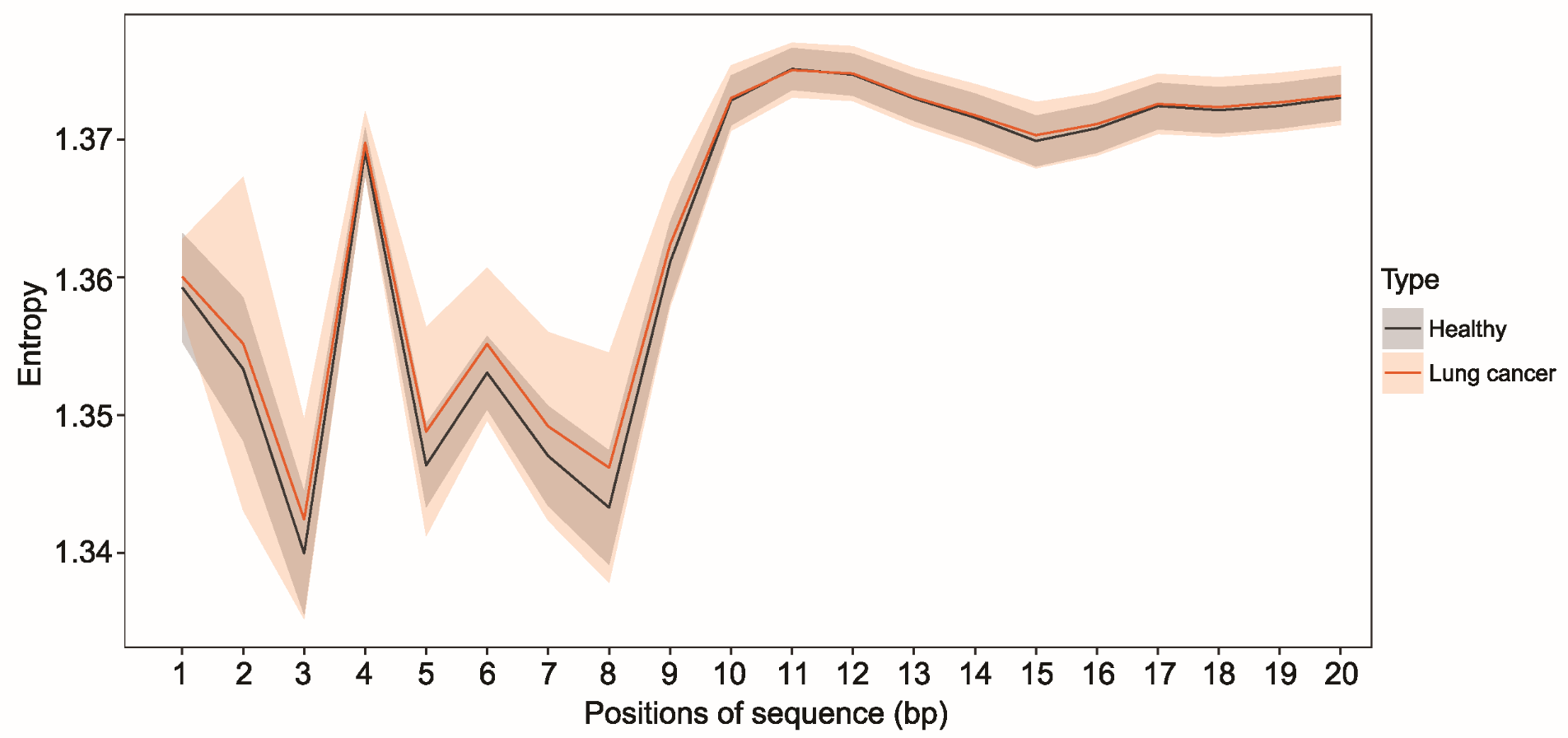

**Fig. S3 Shannon entropy of the first 20 bp at fragment ends**

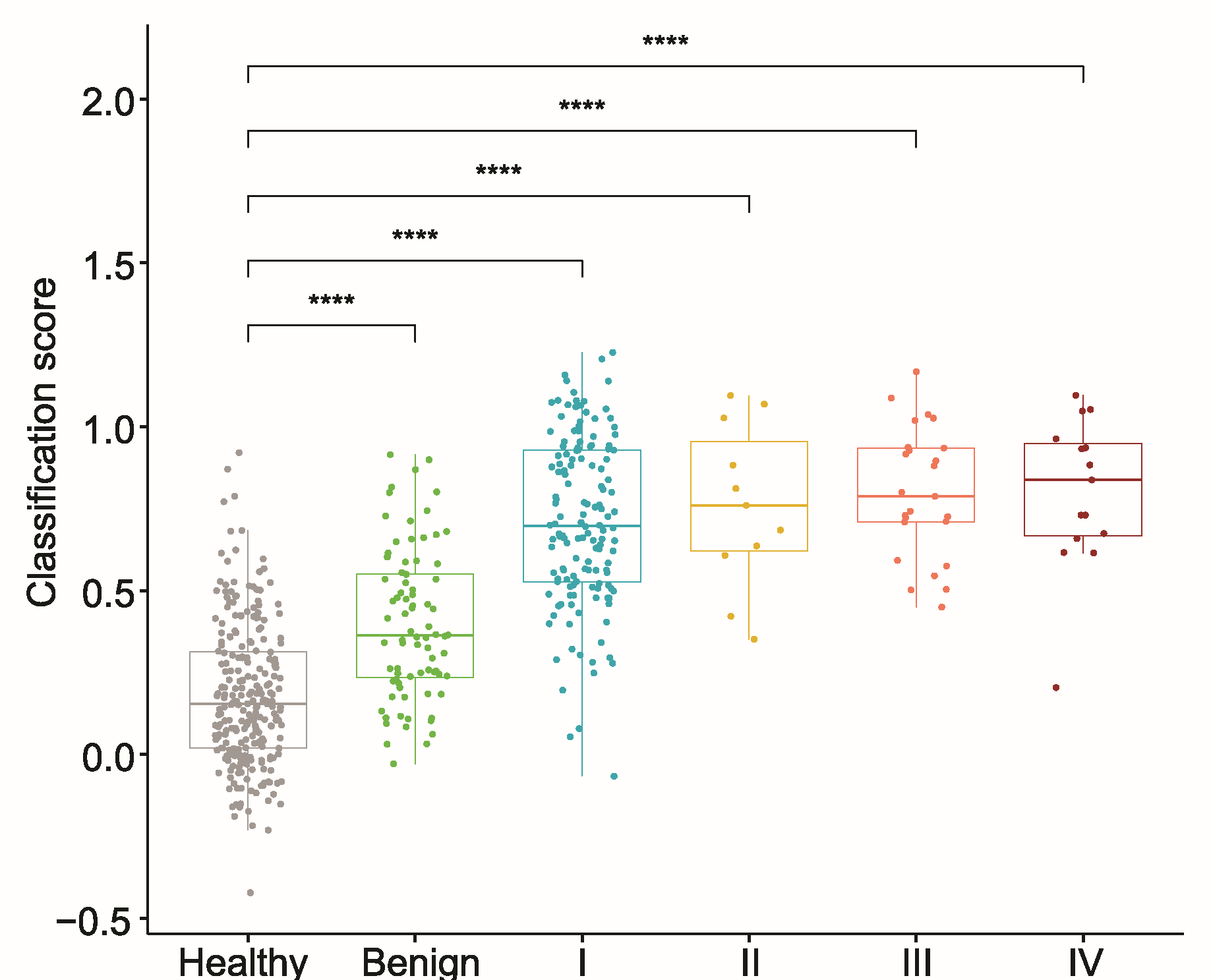

**Fig. S4 Classification score distributions across non-cancer individuals and cancer patients.**

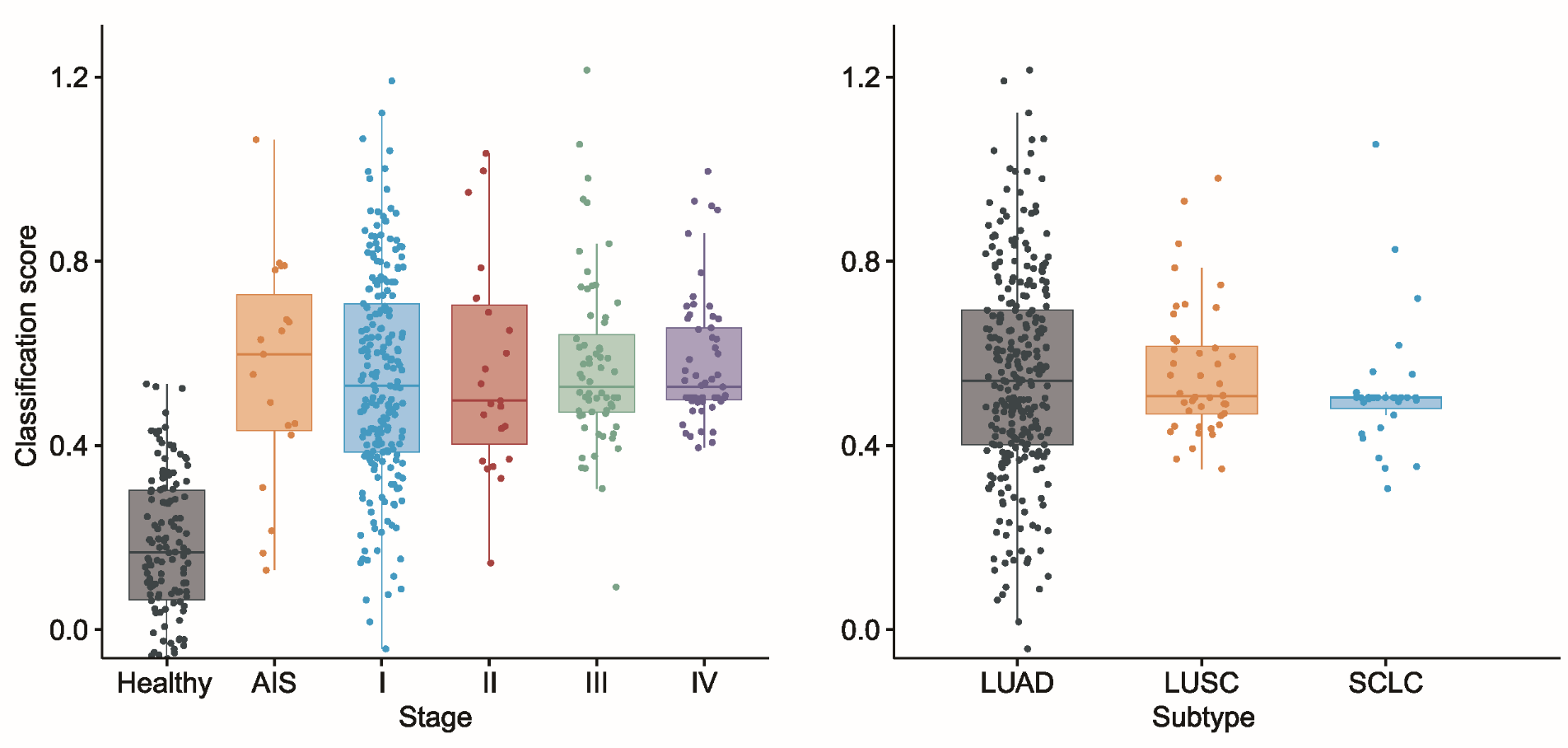

**Fig. S5 Classification score distributions across non-cancer individuals and cancer patients in the independent external test cohort.** (A) Classification score distributions across non-cancer individuals and cancer patients, stratified by clinical stage, and (B) by cancer subtypes.

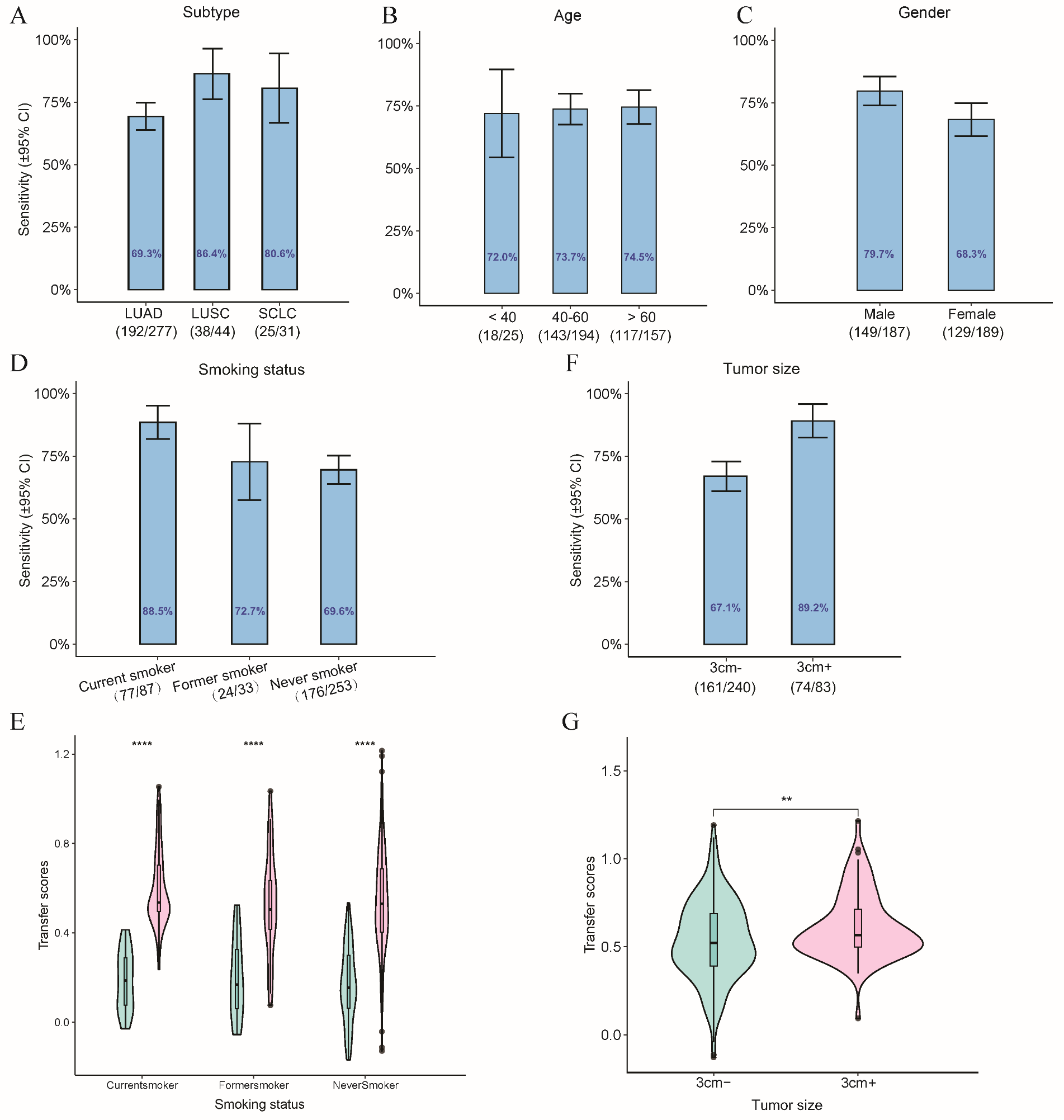

**Fig. S6 Sensitivity of FOTP-based model across different clinical subgroups in the** **independent cohort at 95% specificity.** Histograms show sensitivity with 95% confidence intervals indicated by the bars for various clinical subgroups: (A) cancer subtype, (B) age, and (C) gender. (D) Sensitivity stratified by smoking status and (E) corresponding classification score distributions across healthy individuals and lung cancer patients. (F) Sensitivity by tumor size and (G) corresponding classification score distributions.

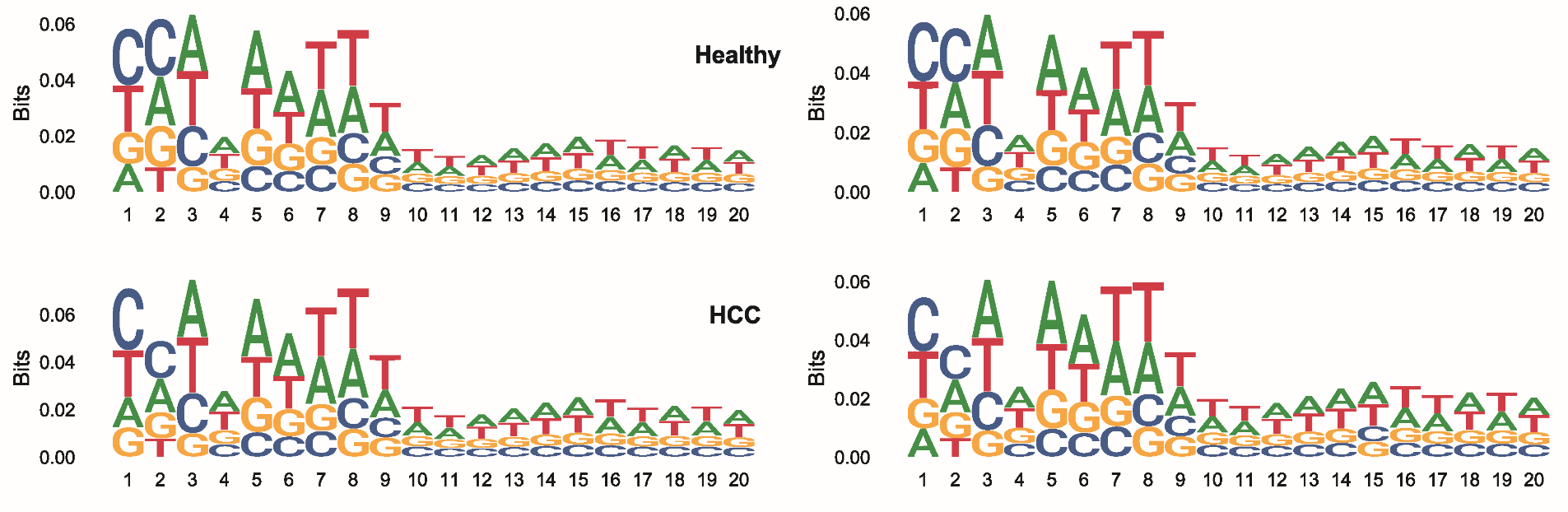

**Fig. S7 Nucleotide frequencies within the first 20 bp from fragment ends for healthy individuals and HCC patients.**
